# Effects of a Program to Reduce Unnecessary Echocardiograms and Nuclear Stress Tests for Cardiopulmonary Complaints in an Emergency Department-Run Observation Unit

**DOI:** 10.64898/2026.09.12.26361477

**Authors:** Maria D’Amico, Diamia Foster, Leslie Tasha Mbapah, Mark Richman

## Abstract

**Introduction:** Inappropriate use of echocardiograms and stress tests with nuclear components in the evaluation of cardiopulmonary complaints (chest pain, dyspnea/shortness of breath (SOB), dizziness, and syncope) is common, despite Cardiology society guidelines for appropriate use. Such overuse raises healthcare costs, impairs the availability of resources for other patients, and exposes patients to radiation unnecessarily. Our Emergency Department (ED) runs an observation unit that sees thousands of patients annually with cardiopulmonary complaints. We observed many have echocardiograms and stress tests with nuclear components without indications for either. Working with our Department of Cardiology, we promoted guidelines for appropriate use of echocardiograms and stress tests with nuclear components in the evaluation of such patients.

**Methods:** We collected 2024 and 2025 data regarding OU admissions. Patents were included if they were ≥18 years, in the ED OU between 1/1/24 and 12/31/25, aad an OU admission complaint of chest pain, dyspnea/SOB, dizziness, and syncope. We calculated the percent of patients with such presentations who received either an echocardiogram or a stress test with a nuclear component. We then compared the 2024 (pre-implementation) data with full-year data from the first full year of implementation (2025).

**Results:** After introducing educational guidelines, there were statistically-significant reductions in nuclear stress tests ordered for dyspnea/SOB (absolute reduction 9.3%; p = 0.0009) and overall usage of nuclear stress tests (absolute reduction 2.4%; p = 0.0095). This was offset by increases in echocardiograms ordered for chest pain (absolute increase 3.7%; p = 0.005) and overall usage of echocardiograms (absolute increase 2.0%; p = 0.0454). There was no net change in the overall use of echocardiograms and nuclear stress tests when considered together: 85.2% of patients had one or the other study in 2024 vs. 84.8% in 2025; p = 0.5768.

**Discussion:** Introducing and promoting appropriate-use guidelines for echocardiograms and stress tests with nuclear components in the evaluation of cardiopulmonary complaints was associated with a reduction in unnecessary nuclear stress tests, but not echocardiograms. Success of this program to reduce nuclear stress tests may be attributed to several factors. First, the relevant specialties (emergency medicine, cardiology) were engaged. Second, we relied on guidelines adopted by professional organizations. Third, the initiative corresponds to nationwide efforts to decrease cost of care by providing evidence-based diagnostic and treatment management. Failure to reduce echocardiograms may be due to a well-documented multi-year time lag between changes in medical knowledge and wide-scale adoption; failure to develop the guidelines in collaboration with house staff and mid-level providers and promote them at multiple times and venues in a multi-modal manner; and utilize Department leadership to promote their usage.

## Introduction

Following initial evaluation, Emergency Department (ED) patients are discharged, admitted, or placed into an observation unit (OU) for further assessment and treatment. The use of OUs has increased over the past 25 years,^1^ as they are associated with decreased length of stay, costs, and increased patient satisfaction.^2^ The Long Island Jewish Medical Center ED staffs a 12-bed OU with attending physician and physician assistant (PA) coverage. The OU has an annual census of ~3,000 patients. Occasionally, when more than 12 patients qualify for observation status, up to two additional patients are put in “observation status” while physically in another part of the ED (“virtual observation”). Patients managed in the OU typically have these conditions: chest pain (for echocardiogram and/or cardiac stress test); mild-moderate asthma; pharyngitis or peritonsillar abscess; transient ischemic attack, minor stroke, or significant back pain needing MRI (which takes a long time in the queue and the machine, and for interpretation); and significant anemia requiring blood transfusion (which takes 3 hours per unit transfused). OU patients are usually kept for 24-36 hours.

After evaluation, ED patients with chest pain are determined to have non-cardiac chest pain, possible acute coronary syndrome (ACS), or definite ACS.^3^ Those with possible ACS are often admitted to an OU. Nationally, 18% of patients admitted to an OU are for complaints for a cardiac evaluation (eg, for chest pain).^4^ A variety of studies (eg. electrocardiograms (EKGs), echocardiograms, non-nuclear stress tests, and nuclear stress tests (which visualize areas where the heart is or is not getting good blood flow)^5,6^) are available to assess heart structure and function, as well as blockages, heart failure, heart attacks, or dysrhythmias.

Absent marked physical examination or EKG abnormalities, the yield of echocardiographic findings leading to immediate, substantial changes in care among patients with cardiopulmonary complaints, including syncope (a common ED OU admission diagnosis), is very low.^7^ Such low utility was demonstrated in a 2019 study of 120 OU patients with syncope.^8^ Only 6.8% of those without physical examination or electrocardiographic findings suggesting a high-likelihood of an actionable echocardiographic abnormality did, indeed, have an actionable echocardiographic abnormality.

In the United States, an estimated eight million nuclear stress tests are performed on patients, many with the indication being “prior myocardial infarction” or “history of coronary artery bypass graft (CABG);”^9^ these, on their own, are not strong indications for adding a nuclear component to a stress test. Reducing unnecessary nuclear stress tests is important for both the health of the patient, as well as cost of care and overuse of medical resources. The average cost of a stress echocardiogram or electrocardiogram is $15-$275, while the price of a nuclear stress test is $400-$750,^5^ depending on a patient’s insurance policy. Aside from the increased cost, a patient who gets a nuclear stress test is also exposed to unnecessary radiation, which can lead to an increased risk of cancer.

The American College of Cardiology (ACC), American Heart Association (AHA), and American Society of Nuclear Cardiology (ASNC) have guidelines for the use of nuclear stress tests, which are most-beneficial in patients with pacemakers, left bundle branch blocks (LBBBs), or who are unable to exercise due to obesity, arthritis, diabetes, or other healthcare issues. Per our OU’s guidelines, indications for the use of nuclear stress testing include patients who have an uninterpretable ECG, a pacemaker or LBBB, or an inability to exercise. Stress tests without a nuclear component are effective for other patient populations. These indications are similar to those listed in the ACC/AHA/ASNC guidelines.

However, we observed that many OU patients with cardiopulmonary complaints are ordered an echocardiogram or stress test with nuclear imaging without clear indication for either. Aligning ordering of echocardiograms and nuclear stress tests with guideline-recommended indications can reduce costs, radiation, and unnecessary resource utilization, contributing to long term health concerns of patients.^10^ This study aimed to improve concordance of ordering echocardiograms and nuclear stress tests with with OU guidelines by educating providers about such guidelines. The primary outcomes of interest were decreases in the percent of OU patients with cardiopulmonary conditions who had an echocardiogram and/or a nuclear stress test.

## Methods

Northwell Health is a 22-hospital health system largely operating in Long Island and New York City. Long Island Jewish Medical Center is a 583-bed tertiary care teaching hospital serving an ethnically and socio-economically diverse population. We reviewed ACC/AHA/ASNC guidelines and collaborated with the LIJ Department of Cardiology. They confirmed the general lack of utility of echocardiograms in observation units when physical examination and EKG do not suggest a significant structural disorder, such as markedly-reduced ejection fraction or valvular disease. We created a guideline reiterating appropriate-use criteria for echocardiograms and nuclear stress tests, which we promoted via a single presentation to house staff, mid-level providers, and faculty physicians at the end of 2024. We also posted the guideline as a 1-page, laminated page at the OU provider desk.

We collected 2024 and 2025 data regarding OU admissions. Patents were included if they were:

- Age ≥18 years
- In the LIJ Adult ED OU between 1/1/24 and 12/31/25
- Had an OU admission complaint for which an echocardiogram or stress test would be a reasonable consideration: chest pain (ICD-10: R07), dyspnea/SOB (ICD-10: R06.00), dizziness (ICD-1): R42), and syncope (ICD-10: R55)

We calculated the percent of patients with such presentations who received either an echocardiogram or a stress test with a nuclear component. We then compared the 2024 (pre-implementation) data with full-year data from the first full year of implementation (2025).

MS Excel 2021 (Microsoft Corporation, Redmond, WA) was used for data management and statistical analyses. MedCalc was used to compare two percentages.^11^ Statistical significance was set a priori at p <0.05.

This research was reviewed and deemed not to meet the definition of research by the Northwell Health Institutional Review Board’s (IRB’s) Human Research Protection Program, indicating that formal IRB approval was not necessary for this study. All observations were compiled in compliance with institutional guidelines for patient privacy and data security. 06.02)

## Results

After introducing educational guidelines, there were statistically-significant reductions in nuclear stress tests ordered for dyspnea/SOB (absolute reduction 9.3%; p = 0.0009) and overall usage of nuclear stress tests (absolute reduction 2.4%; p = 0.0095). This was offset by increases in echocardiograms ordered for chest pain (absolute increase 3.7%; p = 0.005) and overall usage of echocardiograms (absolute increase 2.0%; p = 0.0454). (**Tables 1-5**) There was no net change in the overall use of echocardiograms and nuclear stress tests when considered together: 85.2% of patients had one or the other study in 2024 vs. 84.8% in 2025; p = 0.5768.

**Table 1.** Changes in echocardiograms and nuclear stress tests for chest pain.

|  | 2024 | 2025 | Difference between 2025 and 2024 | P-value |
| --- | --- | --- | --- | --- |
| % of patients with chest pain | 2,898 | 2,849 |  |  |
| Echocardiograms | 1,385 | 1,468 |  |  |
| % with echocardiogram | 47.8% | 51.5% | 3.7% | 0.005 |
| Nuclear stress tests | 1,331 | 1,291 |  |  |
| % with nuclear stress test | 45.9% | 45.3% | -0.6% | 0.648 |

**Table 2.** Changes in echocardiograms and nuclear stress tests for dyspnea-SOB.

|  | 2024 | 2025 | Difference between 2025 and 2024 | P-value |
| --- | --- | --- | --- | --- |
| % of patients with dyspnea/SOB | 426 | 458 |  |  |
| Echocardiograms | 238 | 263 |  |  |
| % with echocardiogram | 55.9% | 57.4% | 1.6% | 0.6531 |
| Nuclear stress tests | 116 | 82 |  |  |
| % with nuclear stress test | 27.2% | 17.9% | -9.3% | 0.0009 |

**Table 3.** Changes in echocardiograms and nuclear stress tests for dizziness.

|  | 2024 | 2025 | Difference between 2025 and 2024 | P-value |
| --- | --- | --- | --- | --- |
| % of patients with dizziness | 729 | 825 |  |  |
| Echocardiograms | 295 | 337 |  |  |
| % with echocardiogram | 40.5% | 40.8% | 0.4% | 0.9044 |
| Nuclear stress tests | 41 | 35 |  |  |
| % with nuclear stress test | 5.6% | 4.2% | -1.4% | 0.2003 |

**Table 4.**
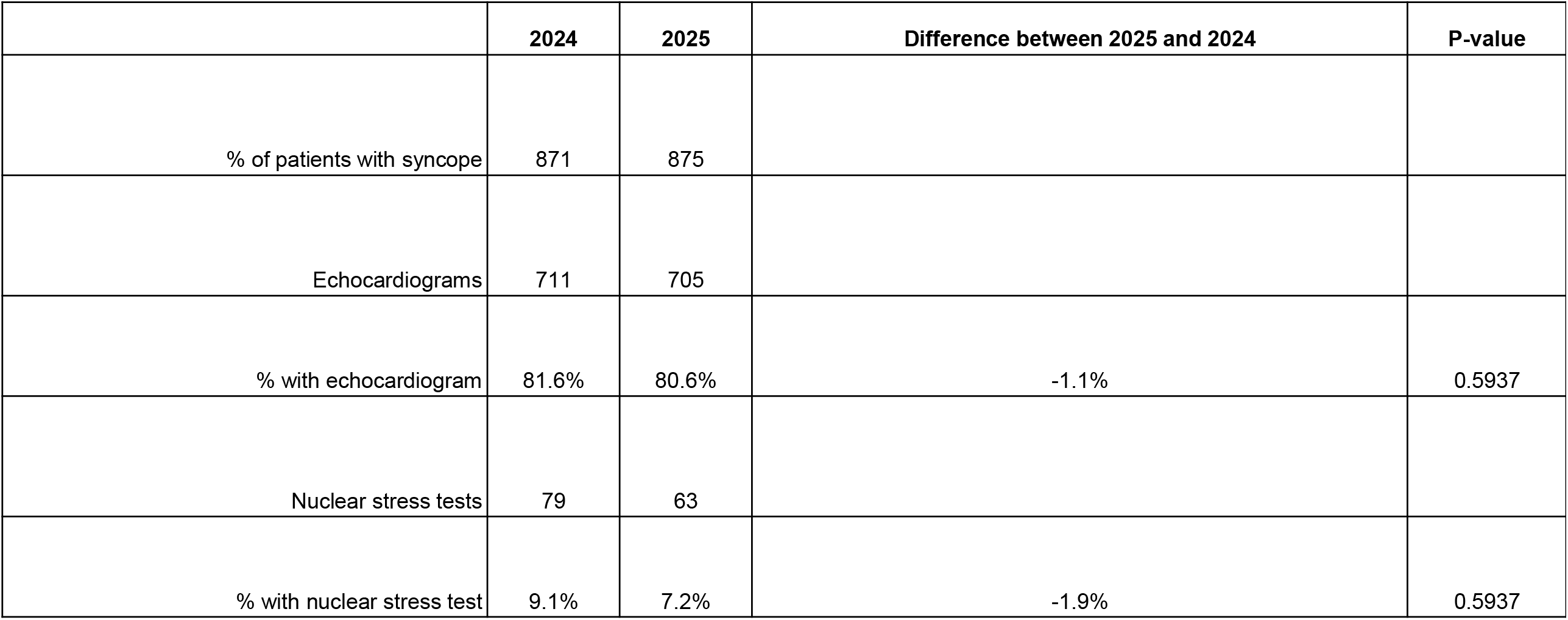
Changes in echocardiograms and nuclear stress tests for syncope.

**Table 5.** Changes in echocardiograms and nuclear stress tests for chest pain, dyspnea-SOB, dizziness, or syncope.

|  | 2024 | 2025 | Difference between 2025 and 2024 | P-value |
| --- | --- | --- | --- | --- |
| # of patients with chest pain, dyspnea/SOB, syncope, or dizziness | 4,924 | 5,007 |  |  |
| Number of echocardiograms | 2,629 | 2,773 |  |  |
| % with echocardiograms | 53.4% | 55.4% | 2.0% | 0.0454 |
| Number of nuclear stress tests | 1,567 | 1,471 |  |  |
| % with nuclear stress test | 31.8% | 29.4% | -2.4% | 0.0095 |

## Discussion

The primary aim of this intervention was to reduce unnecessary observation unit echocardiograms and nuclear component of stress testing, thereby minimizing patient radiation exposure, curbing healthcare costs, and mitigating the overuse of medical resources. Many echocardiograms and nuclear stress tests ordered during cardiac assessments are unnecessary, highlighting the extent of resource overutilization and clinical misuse relative to patient-presenting symptoms. Inappropriate testing places an undue financial burden on both hospitals and patients while needlessly exposing individuals to radiation. This initiative was associated with decreases in nuclear stress test ordering, but not echocardiogram ordering, for chest pain, dyspnea/SOB, dizziness, or syncope. This selective alignment with chief complaints suggests clinicians were more liberal with echocardiograms than with nuclear stress tests, perhaps using the echocardiogram as a “screening” test to determine whether a patient would benefit from the longer, more-expensive, and radiation-exposing nuclear stress test. We did not have an opportunity to investigate this hypothesis by looking at the timing of the orders; for example, “Was a nuclear stress test ordered after an echocardiogram resulted?”

Success of this program to reduce nuclear stress tests may be attributed to several factors. First, the relevant specialties (emergency medicine, cardiology) were engaged. Second, we relied on guidelines adopted by professional organizations. Third, the initiative corresponds to nationwide efforts to decrease cost of care by providing evidence-based diagnostic and treatment management.^12^

On the other hand, we were not successful in decreasing the percent of patients on whom echocardiograms were ordered. This may be due to multiple factors.

First, there is a well-documented multi-year time lag between changes in medical knowledge and wide-scale adoption,^13^ often attributable to the four stages of the awareness-to-adherence model: Awareness, Agreement, Adoption and Adherence.

Second, had we utilized different approaches to guideline development and promotion, this project might have been more effective in achieving its primary aim. Most nuclear test orders are placed by house staff, yet the ACC/AHA/ASNC guidelines were developed primarily by attending physicians. Involving house staff directly in guideline development might have fostered greater buy-in and recall. When guidelines are developed by consensus, by end-users, or by individuals with high perceived credibility, adherence improvements of up to 40% have been demonstrated, largely by instilling a sense of ownership among clinicians.^14^

In addition, employing a multi-modal educational program rather than relying solely on passive announcements would likely have driven broader adoption. Passive dissemination strategies are frequently ineffective, whereas interactive approaches, such as in-person or technology-facilitated workshops, succeed at higher rates. For example, select house staff could have been recruited to introduce the guidelines to peers during educational conferences. In those settings, presenters could use case-based scenarios, polling the audience on whether they would order an echocardiogram or nuclear test for specific cases and discussing appropriate ordering indications.

Another possible hindrance was the frequency of education. We educated ED providers regarding proper test ordering only once. This project may have seen greater success had the educational campaign been repeated several times during 2025. Frequent reminders, audits, and peer review are closely associated with decreases in unnecessary diagnostic testing, and successful reductions in nuclear stress tests have been accomplished when guidelines were paired with hour-long educational sessions and structured reminders.^15^ Involving service-line thought leaders in the reminder process would likely have enhanced these efforts. Effective leadership facilitates successful guideline implementation,^16^ as clinical opinion leaders leverage local networks to exert social influence and drive behavior change. Rather than reviewing all underlying data for a new recommendation, providers frequently rely on guidance from key peer opinions when deciding whether to follow clinical guidelines. Clinical opinion leaders help diffuse criticism, identify and overcome barriers, and operationalize quality initiatives. In our initiative, leadership figures within the LIJ ED and cardiology services could have organized follow-up communications, digital education, and regular reminders.

Ultimately, the decision to adhere to a guideline depends on several factors: ease of use, applicability to the patient population, physician autonomy, institutional culture and common practice, point-of-care reminders, and perceived legal risk. However, had we adopted the elements described above as part of this initiative, the initiative may have been even more effective, perhaps yielding a reduction in echocardiograms in addition to a reduction in nuclear stress tests.

### Limitations

This study has several limitations. First, as a quality improvement project (rather than formal research), our findings cannot be extrapolated to other settings, even those similar to ours. In addition, it was a single-site study, and there was a historical control group (2024 vs. 2025) rather than a randomized control group as part of a randomized, controlled trial (RCT). Consequently, pre-vs. post-implementation patients may not have been similar in ways that influenced whether they had a nuclear stress test or echocardiogram. Conducting an RCT in this situation would have been logistically difficult, as it would have required exposing half of the providers to the intervention (the echocardiogram and nuclear stress test guidelines) while not exposing the other half. Since physicians and mid-level providers work with different colleagues during each ED shift, and share knowledge and practice patterns with one another, there would inevitably have been a mix on most shifts between providers in the intervention group and those in the control group, thereby contaminating the effect of the intervention. Furthermore, we did not review each record to determine whether, in any particular OU patient, the echocardiogram or nuclear stress test was indicated or not according to our OU’s guidelines. For example, an echocardiogram in a particular patient might have been indicated as likely to lead to a change in management (such as if there were a new-onset significant murmur in the setting of established congestive heart failure) or a nuclear component of a stress test might have been appropriate in a patient with a LBBB. It is possible some of the echocardiograms and nuclear stress tests were ordered on such patients, and, therefore, would have been indicated and not “unnecessary.” Finally, some echocardiograms and nuclear stress tests may have been indicated by a change in the patient’s condition during their OU stay.

## Data Availability

All data produced in the present work are contained in the manuscript

